# SARS-CoV-2 virologic rebound with nirmatrelvir-ritonavir therapy

**DOI:** 10.1101/2023.06.23.23288598

**Authors:** Gregory E. Edelstein, Julie Boucau, Rockib Uddin, Caitlin Marino, May Y. Liew, Mamadou Barry, Manish C. Choudhary, Rebecca F. Gilbert, Zahra Reynolds, Yijia Li, Dessie Tien, Shruti Sagar, Tammy D. Vyas, Yumeko Kawano, Jeffrey A. Sparks, Sarah P. Hammond, Zachary Wallace, Jatin M. Vyas, Amy K. Barczak, Jacob E. Lemieux, Jonathan Z. Li, Mark J. Siedner

## Abstract

**Objective:** To compare the frequency of replication-competent virologic rebound with and without nirmatrelvir-ritonavir treatment for acute COVID-19. Secondary aims were to estimate the validity of symptoms to detect rebound and the incidence of emergent nirmatrelvir-resistance mutations after rebound.

**Design:** Observational cohort study.

**Setting:** Multicenter healthcare system in Boston, Massachusetts.

**Participants:** We enrolled ambulatory adults with a positive COVID-19 test and/or a prescription for nirmatrelvir-ritonavir.

**Exposures:** Receipt of 5 days of nirmatrelvir-ritonavir treatment versus no COVID-19 therapy.

**Main Outcome and Measures:** The primary outcome was COVID-19 virologic rebound, defined as either (1) a positive SARS-CoV-2 viral culture following a prior negative culture or (2) two consecutive viral loads ≥4.0 log_10_ copies/milliliter after a prior reduction in viral load to <4.0 log_10_ copies/milliliter.

**Results:** Compared with untreated individuals (n=55), those taking nirmatrelvir-ritonavir (n=72) were older, received more COVID-19 vaccinations, and were more commonly immunosuppressed. Fifteen individuals (20.8%) taking nirmatrelvir-ritonavir experienced virologic rebound versus one (1.8%) of the untreated (absolute difference 19.0% [95%CI 9.0-29.0%], P=0.001). In multivariable models, only N-R was associated with VR (AOR 10.02, 95%CI 1.13-88.74). VR occurred more commonly among those with earlier nirmatrelvir-ritonavir initiation (29.0%, 16.7% and 0% when initiated days 0, 1, and ≥2 after diagnosis, respectively, P=0.089). Among participants on N-R, those experiencing rebound had prolonged shedding of replication-competent virus compared to those that did not rebound (median: 14 vs 3 days). Only 8/16 with virologic rebound reported worsening symptoms (50%, 95%CI 25%-75%); 2 were completely asymptomatic. We detected no post-rebound nirmatrelvir-resistance mutations in the NSP5 protease gene.

**Conclusions and Relevance:** Virologic rebound occurred in approximately one in five people taking nirmatrelvir-ritonavir and often occurred without worsening symptoms. Because it is associated with replication-competent viral shedding, close monitoring and potential isolation of those who rebound should be considered.

## Introduction

Data are conflicting about whether nirmatrelvir-ritonavir (N-R) is associated with virologic rebound (VR).^1–7^ However, precise estimation of VR incidence with and without N-R use has been limited by infrequent and short-term sampling, symptomatic reporting, and absence of culture data.

## Methods

The Post-vaccination Viral Characteristics Study (POSITIVES) is a prospective, observational cohort of individuals with acute COVID-19 with longitudinal sampling for viral load, viral culture, and symptom reporting (**supplementary appendix**).^8,9^ Participants are sampled from automated medical record reports in the Mass General Brigham healthcare system on individuals with positive testing or a prescription for COVID-19 therapeutics.

Participants self-collect anterior nasal swabs three times a week for two weeks and weekly thereafter until SARS-CoV-2 viral load testing is persistently undetectable. Specimens are analyzed for SARS-CoV-2 viral load, viral culture, and whole genome sequencing. Participants complete 10-item COVID-19 symptom surveys, graded as absent (0), mild (1), moderate (2), or severe (3), for a maximum total symptom score (TSS) of 30-points. Study physicians complete chart reviews to determine COVID-19 vaccination and treatment history, and immunosuppression status (**STable1)**.

We sought to estimate the incidence of virologic rebound, which we defined in individuals with either 1) positive SARS-CoV-2 viral culture following a negative culture or 2) a viral load ≥1.0 log_10_ from a prior viral load and ≥4.0 log_10_ copies/mL for two consecutive timepoints after a prior reduction in viral load to <4.0 log_10_ copies/mL. We selected this outcome as a surrogate for putative transmission risk, based on data relating transmission to replication-competent virus with viral loads >4.0 log_10_ copies/mL.^10,11^ For a secondary outcome, we redefined VR as a viral load at days 10 and 14 ≥2.7 log_10_ and at least 0.5 log_10_ greater than the result at day 5, in order to compare our estimates to the EPIC-HR study, which considered fewer time points and did not incorporate culture methods.^1^

Our primary exposure of interest was exposure to N-R therapy. Therefore, we limited analysis to ambulatory participants enrolled after March 2022, when we began recruiting individuals initiating N-R. We also excluded participants without a nasal swab collected >11 days from their first positive COVID-19 test, because approximately 90% of rebound phenomena occur by this time,^8^ and individuals who received N-R for more or less than 5 days. We compared the frequency of VR by N-R use overall and stratified by potential confounders (i.e., immunosuppression, age, sex, and prior COVID-19 vaccinations) using two-sided Fisher’s exact tests, and after adjustment for confounders, in logistic regression models. We compared the frequency of VR by timing of N-R initiation, using a non-parametric test of trend. We compared our estimate of VR with the definition used in the EPIC-HR study.^1^ We used the Kaplan-Meier survival estimator to depict and compare days to initial and final viral culture negativity, stratified by N-R use and VR, using log-rank testing. We assessed the validity of symptom rebound, as defined by an increase in TSS by 3 or more points from a prior date, and the presence of any symptoms during the rebound period, to detect VR.^6^ Finally, we report the proportion of sequenced viruses before and after VR with mutations in the NSP5 gene encoding the main protease (M^pro^) of SARS-CoV-2. Statistical analyses and figure production were conducted with Stata version 16.1 and GraphPad Prism version 9.5.

### Ethical Considerations

All study participants provided verbal informed consent. Written consent was waived by the ethics committee, based on the involvement of participants with acute COVID-19 in a minimal risk study. The study procedures were approved by Institutional Review Board and the Institutional Biosafety Committee at Mass General Brigham.

## Results

Compared with untreated individuals (n=55), those taking N-R (n=72) were older (57 vs 39 years, P<0.001), received more COVID-19 vaccinations (median 4 vs 3, P<0.001) and were more commonly immunosuppressed (32% vs 9%, P<0.001, **SFig1/STable2**). Fifteen individuals (20.8%) taking N-R experienced VR versus one (1.8%) untreated individual (**Figures 1&2**, absolute difference 19.0% [95%CI 9.0-29.0%], P=0.001). In sub-group analyses, VR was numerically more frequent in all demographic and clinical sub-groups (**Figure 2**). In multivariable logistic regression models including demographic and clinical characteristics, only N-R use remained associated with VR (**STable 3**). There was a trend towards higher rates of VR with earlier N-R initiation (29%, 16.7% and 0% when initiated days 0, 1, and ≥2 after diagnosis, P=0.089, **Figure 2**). When we restricted analyses to three timepoints, as done in the EPIC-HR study, only 3/124 (2.4%) had rebound detected, and 13/16 (81.2%) rebound events were not captured (**Figure 1E-F**). We detected no post-N-R drug resistance mutations in the NSP5 protease gene (**SFig2**).

**Figure 1.**
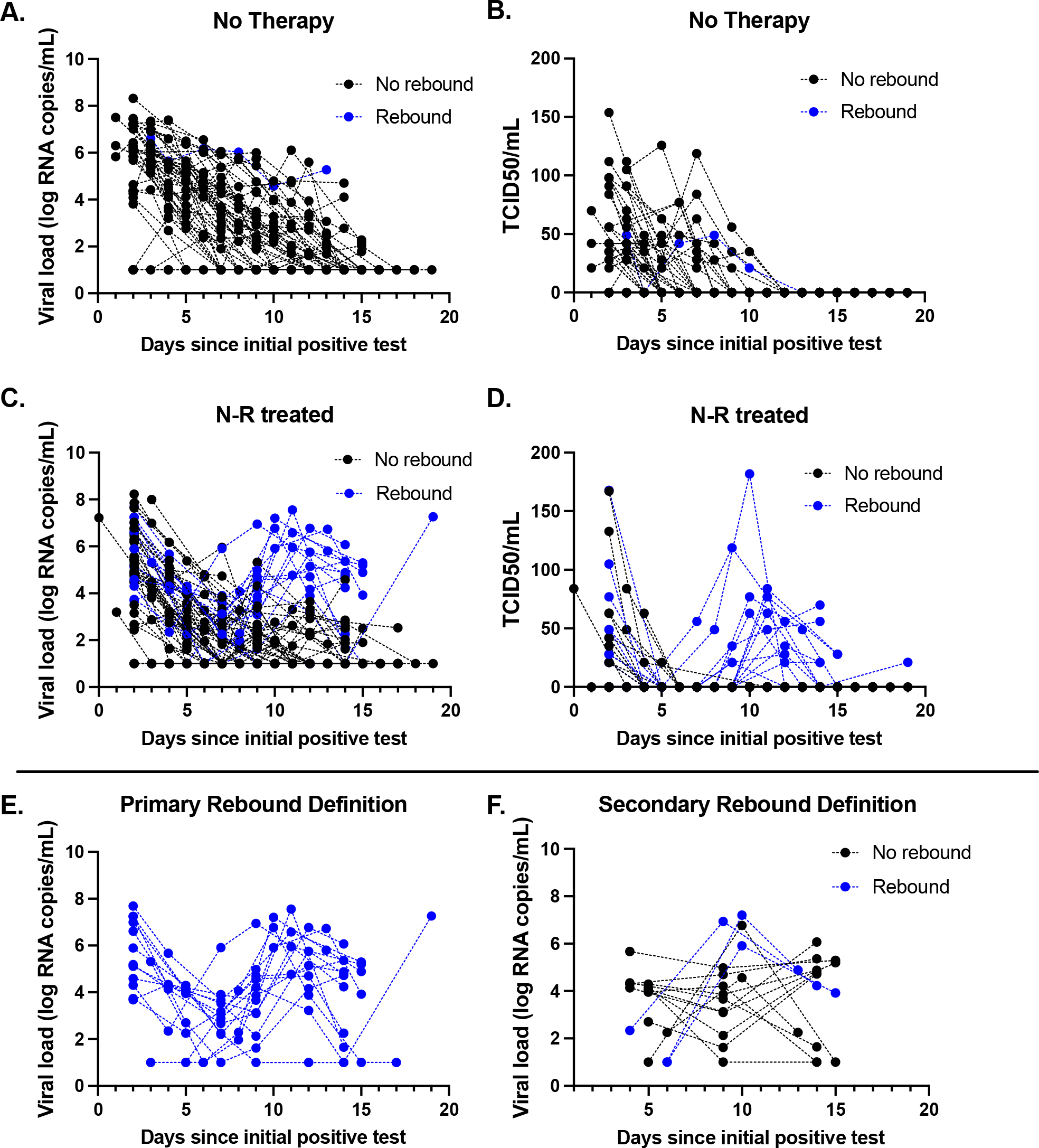
Virologic decay curves with semiquantitative viral cultures and quantitative viral load among individuals with acute COVID-19 taking no therapy or nirmatrelvir-ritonavir (N-R). Black lines indicate individuals without rebound, whereas blue lines indicate individuals with virologic rebound. Panels A (viral load) and B (viral culture) depict decay curves for those not receiving therapy. Panels C (viral load) and D (viral culture) depict individuals who received N-R. Panels E and F compare our primary outcome with all available time points (E) or restricted to days 5, 10 and 14 only (Panel F) as defined in prior studies [1]. Using only three timepoints to detect rebound resulted in missing 81% of the observed virologic rebound events of replication-competent virus.

**Figure 2.**
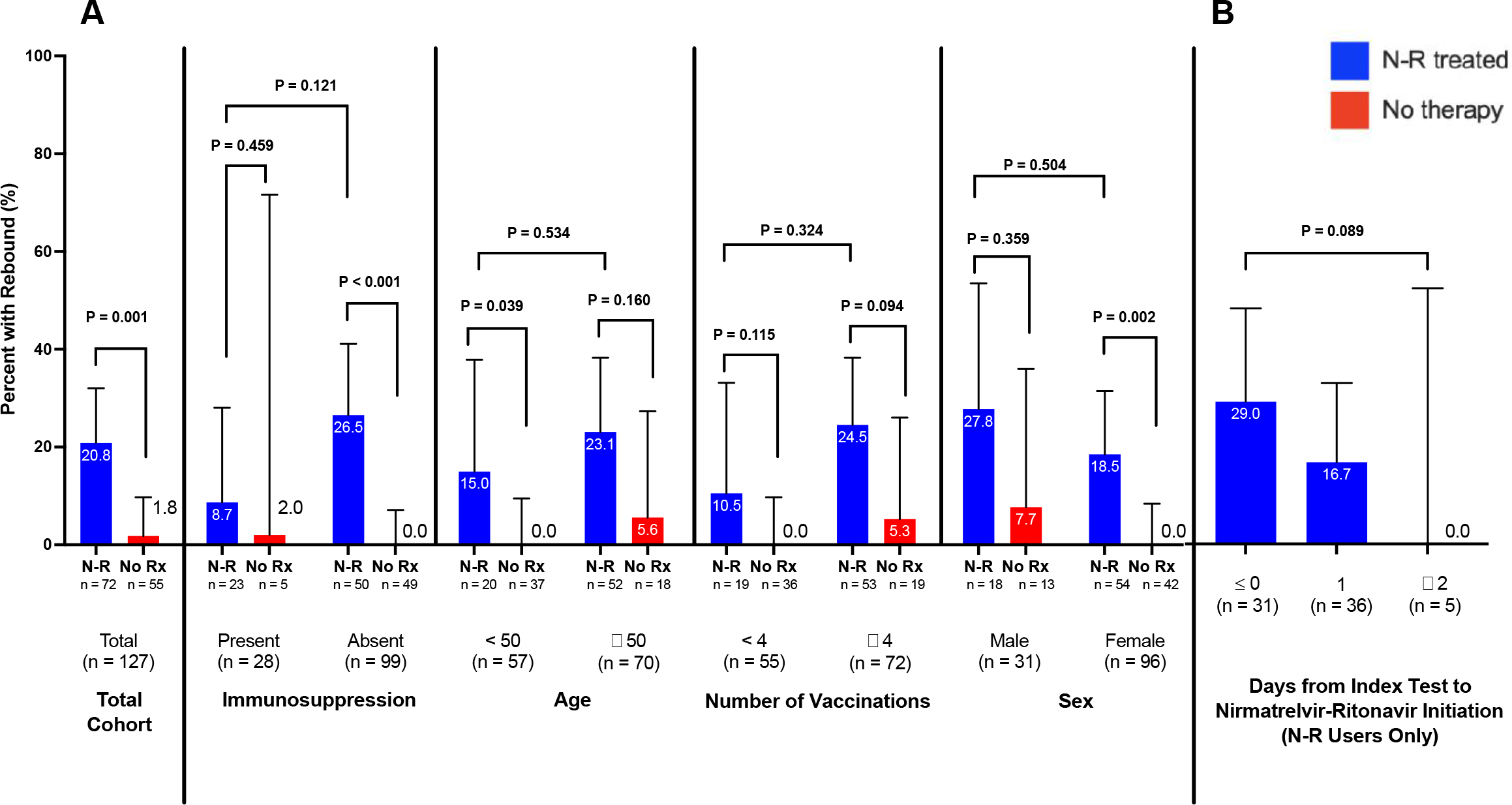
Comparative frequency of virologic rebound by nirmatrelvir-ritonavir use, stratified by demographics and clinical characteristics (A), and by number of days between the first positive SARS-CoV-2 test and initiation of nirmatrelvir-ritonavir therapy (B). For the sub-group comparisons, the bottom P-values represent Fisher’s exact tests comparing rebound rates between those taking versus those not taking nirmatrelvir-ritonavir. The upper P-values represent Fisher’s exact tests comparing rebound rates among those taking nirmatrelvir-ritonavir across the sub-groups, for example comparing those taking nirmatrelvir-ritonavir with immunosuppression present versus those taking nirmatrelvir-ritonavir with immunosuppression absent.

N-R recipients achieved initial culture conversion sooner than those not treated (**Figure 3/STable4**, P<0.001). However, days to final culture conversion was similar (**Figure 3**, P=0.29) because those experiencing VR had significantly prolonged shedding (median 14 [IQR13-20] vs 3 days [IQR2-4], **Figure 3/STable4/STable5**). Only 8/16 with VR reported symptom rebound (50%, 95%CI 25-75%); 2 were totally asymptomatic. Only 8/27 with symptom rebound had VR (30%, 95% CI 14-50%, **SFig3/STable6**).

**Figure 3.**
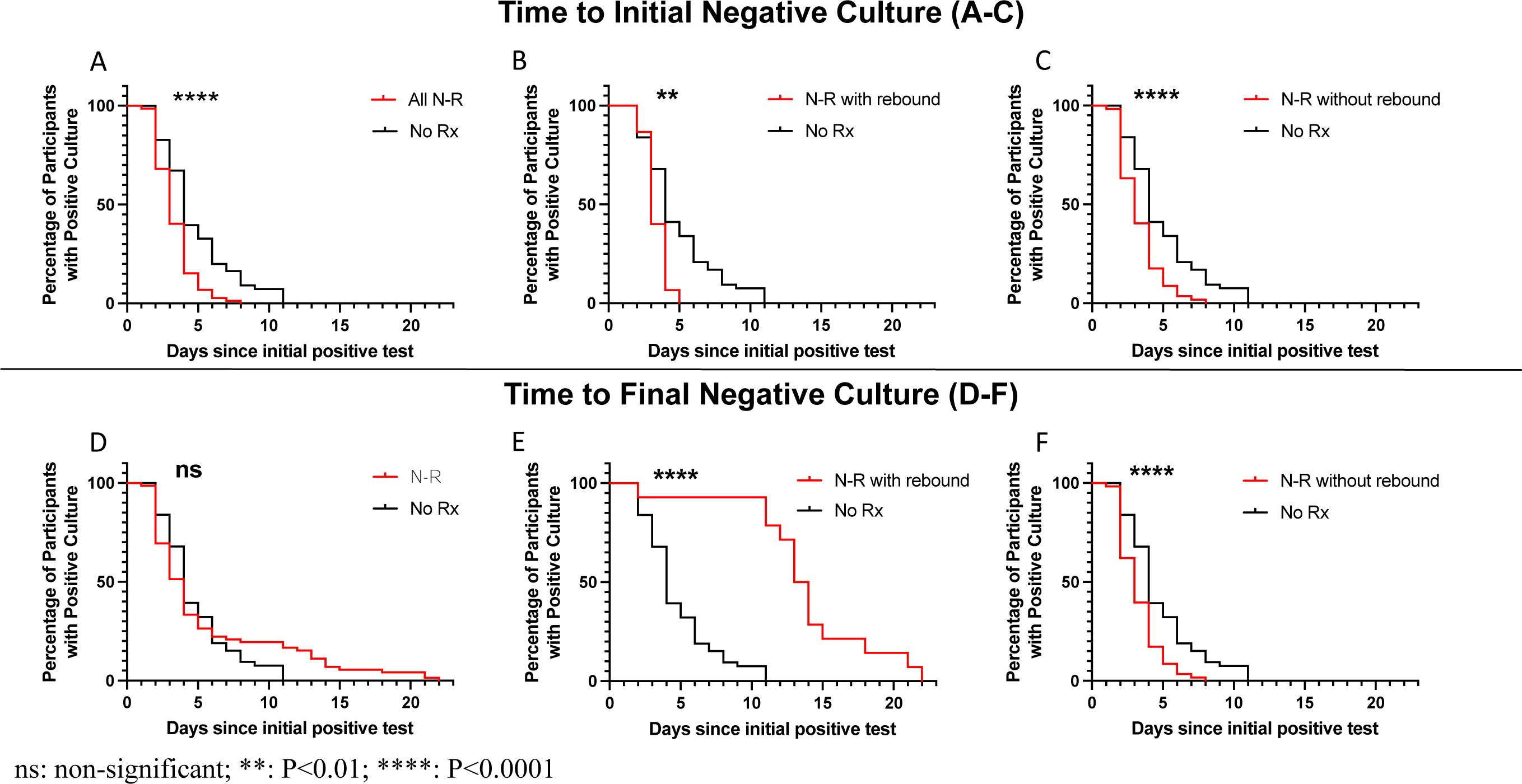
Kaplan Meier survival curves demonstrating time from initial positive SARS-CoV-2 test until initial negative viral culture (A-C) and final negative culture (D-F). In Panel A, we demonstrate that there is a faster time to first negative culture in those receiving nirmatrelvir-ritonavir (N-R) versus no therapy (No Rx). In Panels B and C, we find similar patterns in time to initial negative culture, when dividing the N-R group into those who rebounded (B) and those who did not (C). However, as shown in Panel D, there is no difference in time to final negative culture between N-R and No Rx groups. This appears to be due to the prolonged time to final negative culture among N-R users who rebound (Panel E), because the time to final negative culture remains shorter in N-R users who did not rebound compared to the No Rx group (Panel F).

## Discussion

VR with replication-competent viral shedding occurred in approximately 20% of those taking N-R and 2% of those not on therapy. N-R use remained associated with VR after adjustment for demographic and clinical characteristics, such as vaccination and immunosuppression status. Although N-R treated individuals took fewer days to achieve initial culture negativity, time to final culture negativity was similar, due to prolonged shedding of replication-competent virus among those experiencing VR (median 14 vs 3 days). These data support the presence of an N-R-associated virologic rebound phenomenon, which substantially increases the duration of shedding of replication-competent virus and has implications for post-N-R monitoring and isolation recommendations.

We found a higher incidence of VR with N-R use than prior studies. We believe this is due to use of frequent sampling and culture methods to detect VR. When we restricted our analysis to three PCR-based timepoints, as done in prior trials,^1^ we detected a 2.4% rate of VR, which approximates prior studies, but notably missed 80% of VR events.

VR appeared to be less common among those who delayed therapy by 1 or 2 days after their first positive test. This finding, in conjunction with the lack of drug resistance-associated mutations after VR events, promotes hypotheses that VR may occur due to incomplete viral eradication,^12^ and supports studies to evaluate longer durations of N-R therapy.^13^

Finally, symptoms should not be relied upon to detect or exclude VR. Two individuals with VR had a complete absence of symptoms during the VR period and less than half had symptom rebound. Conversely, the majority of those who did have symptom rebound did not experience VR.

Our study was limited by an observational design, with expected differences between those taking N-R and untreated individuals based on treatment guidelines for N-R^14^. Nonetheless, VR remained associated with N-R, even after adjustment for potential confounders. We used viral culture as a surrogate for transmission risk but did not measure contagiousness or transmission events directly.

These data support a relationship between N-R use and VR. Future work should elucidate the mechanistic pathways of VR, determine if delays in initiating N-R or longer courses of N-R may prevent VR among high-risk individuals, and evaluate larger samples to identify the risk factors for N-R-associated VR.

## Data Availability

All data produced in the present study are available upon reasonable request to the authors

## Acknowledgments

We would like to thank the study participants for their time and considerable efforts to provide specimens in the acute phase of illness as part of this project.

## Notes

**Funding:** This work was supported by the National Institutes of Health (U19 AI110818), the Massachusetts Consortium for Pathogen Readiness SARS-CoV-2 Variants Program and the MGH Department of Medicine. Additional support was provided by the Ragon Institute BSL3 core, which is supported by the NIH-funded Harvard University Center for AIDS Research (P30 AI060354). Drs. Sparks and Wallace are supported by the National Institute of Arthritis and Musculoskeletal and Skin Diseases (R01 AR080659). Dr. Sparks is also supported by the Llura Gund Award for Rheumatoid Arthritis Research and Care. The funders had no role in study design; in the collection, analysis, and interpretation of data; in the writing of the manuscript; or in the decision to submit the manuscript for publication.

### Competing Interest Statement

AKB reports consulting for ICON Government and Public Health Solutions. JZL reports consulting for Abbvie and research funding from Merck. SPH reports research funding from GlaxoSmithKline and has served on an advisory board for Pfizer.

### Funding Statement

This work was supported by the National Institutes of Health (U19 AI110818), the Massachusetts Consortium for Pathogen Readiness SARS-CoV-2 Variants Program and the MGH Department of Medicine. Drs. Sparks and Wallace are supported by the National Institute of Arthritis and Musculoskeletal and Skin Diseases (R01 AR080659). Dr. Sparks is also supported by the Llura Gund Award for Rheumatoid Arthritis Research and Care. The funders had no role in study design; in the collection, analysis, and interpretation of data; in the writing of the manuscript; or in the decision to submit the manuscript for publication.

### Author Declarations

Study procedures were approved by the human subjects review committee at Mass General Brigham and all participants gave informed consent to participate.

