## Supplementary Appendix for "SARS-CoV-2 virologic rebound with nirmatrelvir-ritonavir therapy"

This appendix has been provided by the authors to give readers additional information about their work.

**<u>Supplement to</u>**:

Supplement Version Date: 23 June 2023

### Appendix Table of Contents

**Author Contributions**

**Study Conception and Design:** A.K.B., J.B., J.E.L., J.Z.L., and M.J.S.

**Data Collection**: G.E.E., J.B., R.U., C.M., M.Y.L., M.B., M.C.C., R.F.G, Z.R., Y.L., D.T., S.S., T.D.V., Y.K., J.A.S., S.P.H., Z.S.W., J.M.V

**Analysis and Interpretation of Results:** G.E.E., J.B., R.U., M.C.C., A.K.B, J.E.L., J.Z.L, M.J.S.

**Draft Manuscript Preparation:** G.E.E., M.J.S.

**Provided editorial input to the manuscript and approved the final version:** All authors

### Supplementary Methods

*Study Design*

POSITIVES is an observational cohort study that enrolls adults (age ≥18) who test positive for COVID-19 in the Mass General Brigham healthcare system (Boston, Massachusetts, USA).[1,2] An automated list of individuals in the health system with a positive test and/or a prescription for a COVID-19 therapeutic is used as a recruitment frame. Participants may also self-refer from online study information sheets or be referred by healthcare providers. Consenting participants undergo an initial medical chart review by study physicians to determine their COVID-19 vaccination status, treatment history and medical history. Individuals with a diagnosis of leukemia or lymphoma, those with a history of solid organ or bone marrow transplant, and those receiving immunosuppressive therapies including corticosteroids, interferon-gamma inhibitors, or cytotoxic therapies (e.g., anti-cytokine therapies) are classified as being immunosuppressed (**Supplemental Table S1**).

*Study Procedures*

Starting at enrollment, participants self-collect anterior nasal swabs approximately three times a week for two weeks and weekly thereafter until SARS-CoV-2 viral load testing is persistently undetectable. On each date of swab collection, participants complete a 10-item acute COVID-19 symptom survey, with each graded as absent (0 points), mild (1 point), moderate (2 points), or severe (3 points), allowing for a maximum total symptom score (TSS) of 30-points.

*Quantitative Viral Load Assay*

Quantification of SARS-CoV-2 viral load was performed as previously described.[3] Briefly, anterior nasal swabs were placed in viral transport media (VTM), which was then aliquoted in 250 µL. 10 µL of replication-competent avian retrovirus (RCAS) virion was added to each sample as an internal quality control, and the homogenized mixtures were pelleted at 21,000 x g for 2 hours at 4$^oC$. The supernatant was discarded, and 750 µL TRIzol-LS Reagent (ThermoFisher Scientific) was added and vortexed for 30 seconds. Following incubation on ice for 10 minutes, 200 µL of chloroform (MilliporeSigma) was added, and the mixtures were vortexed for 30 seconds. Phase separation was accomplished via centrifugation at 21,000 x g for 15 minutes at 4$^oC$. The aqueous RNA-containing layer was isolated and added to tubes containing 100 µL 3 M Sodium Acetate (Life Technologies) and 1.5 µL GlycoBlue Co-precipitant (ThermoFisher Scientific). 300 µL of Isopropanol (MilliporeSigma) was added and the mixtures were shaken, incubated in dry ice for 15 minutes, and then centrifuged at 21,000 x g for 45 minutes at 4$^oC$ to precipitate RNA pellets. Afterwards the supernatant was discarded, and RNA pellets were washed with 900 µL cold 70% ethanol. RNA pellets were resuspended in diethylprocarbonate-treated Water (ThermoFisher Scientific) and used for RT-qPCR with the US CDC 2019-nCoV_N1 primer and probe set (Integrated DNA Technologies). Absolute quantification of viral load was achieved via comparison to a standard curve generated by a 16-fold serial dilution of N1 RNA run on the same plate. All plates contained two non-template control wells and a positive and negative control for N1. The efficiency of the RNA extraction and RT-qPCR amplification was evaluated by quantifying the RCAS RNA recovered from each sample and the two N1 controls. The importin-8 (IPO8) human housekeeping gene was also amplified and evaluated as a measure of sample collection quality. Samples were run in triplicate wells for N1, and in duplicate wells for RCAS and IPO8.

*Viral Culture*

Semi-quantitative viral culture was performed in the BSL3 laboratory of the Ragon Institute of MGH, MIT, and Harvard as previously reported.[1] Vero-E6 cells (ATCC) were maintained in DMEM (Corning) supplemented with HEPES (Corning), 1X Penicillin/Streptomycin (Corning), 1X Glutamine (Glutamax, ThermoFisher Scientific), and 10% Fetal Bovine serum (FBS) (Sigma), harvested using Trypsin-EDTA (Fisher Scientific) and plated at 20,000 cells per well in 96w plates 16-20 hours before infection. Aliquoted VTM specimens were thawed on ice and filtered through either Spin-X 0.45µm or 0.65 µm filters (Corning) at 10,000 x g for 5 minutes. 2 µL of the undiluted filtrate was added to four wells of a 96w plate and serially diluted (1:5) in media containing 5 µg/milliliter (mL) of polybrene (Santa Cruz Biotechnology) before spinfection for 1 hour at 2000 x g at 37$^{o}$C. Each 96w plate contained wells inoculated with SARS-CoV-2 isolate USA-WA1/2020 strain (BEI Resources) as a positive control and medium only as a negative control. The viral culture plates were scored 7 days post-infection by observation under a light microscope and wells showing cytopathic effect (CPE) counted as positive. A median tissue culture infectious dose (TCID50) was calculated using the Spearman-Karber method. For each well showing CPE, the culture supernatant was harvested for virus expansion and RNA isolation using QIAamp Viral RNA Mini kit (QIAGEN) for confirmation of the viral sequence.

*SARS-CoV-2 Whole Genome Sequencing*

Whole genome sequencing was carried out using the Illumina COVIDseq Test protocol as previously described.[2] Briefly, DNA libraries were constructed using the Illumina COVIDSeq

Test Kit, pooled together, and then quantified with a Qubit High Sensitivity dsDNA kit (Invitrogen). Afterwards, genomic sequencing was performed on an Illumina NextSeq 2000 instrument. Sequenced genomes were demultiplexed and assembled on the Terra platform (app.terra.bio). Complete genomes (sequence assembly length greater than 24000 base pairs) were assigned a Pango lineage (https://github.com/cov-lineages/pangolin-data) and deposited to NCBI GenBank under Project Accession PRJNA759255.

*Outcomes*

Our primary outcome was virologic rebound, which we defined in individuals with either 1) a positive SARS-CoV-2 viral culture following a prior negative culture or 2) sustained elevated viral load, characterized by the combination: a) a nadir viral load $< 4.0 \log_{10}$ copies/ mL followed by a viral load $\geq 1.0 \log_{10}$ greater than the nadir; and b) two consecutive viral load results of $\geq 4.0 \log_{10}$ copies/mL. We selected this primary outcome as a surrogate for putative transmission risk, based on prior data relating transmission risk and replication-competent virus with viral loads $\geq 4.0 \log_{10}$ copies/mL.[4,5] For a secondary outcome, we restricted the cohort to viral load measurements at days 5, 10 and 14 (all ±1 day) and defined virologic rebound, as done in the secondary analysis of the EPIC-HR nirmatrelvir-ritonavir phase 3 trial, [6] when viral load days 10 and 14 was $\geq 2.7 \log_{10}$ and at least $0.5 \log_{10}$ greater than the result at day 5. If only day 10 or day 14 viral load data were available, a single measurement on that day $\geq 2.7 \log_{10}$ and at least 0.5 $\log_{10}$ greater than the result at day 5 also met criteria. Individuals missing either day 5 or both day 10 and 14 viral loads (n=3) were excluded from this analysis. We selected this outcome to enable comparison of our results with prior studies and to determine if the additional sampling done in our study enabled increased detection of rebound events.

*Statistical Analysis*

We limited this analysis to ambulatory participants who were enrolled after March 2022, when we began recruiting individuals at the time of N-R initiation. We excluded participants without at least one nasal swab collected on or after day 12 from their first positive COVID-19 test, because approximately 90% of rebound phenomena occur by this time.[2] We divided the cohort into two groups: 1) those receiving N-R therapy and 2) those not receiving N-R therapy. We excluded individuals receiving N-R therapy who did not have a nasal swab collected within one day of completion of N-R therapy, to avoid enrollment of individuals experiencing rebound at the time of study initiation. We also excluded individuals who received N-R for less or more than 5 days and those in either group who received alternate antiviral therapies (i.e., remdesivir, molnupiravir, or monoclonal antibodies).

We graphically depicted virologic decay curves, stratified into N-R use and no therapy groups. We then compared the crude frequency of virologic rebound by N-R use, and stratified by the presence or absence of immunosuppression, age (< versus ≥50 years), sex, and number of prior COVID-19 vaccinations (< versus ≥ 4 prior vaccinations) using two-sided Fisher's exact tests. To assess for confounding, we fit logistic regression models, with VR as the dependent variable, and each of the above demographic and clinical characteristics as independent variables, both alone and in a fully adjusted multivariable model. To compare virologic rebound frequency by timing of initiation of N-R, we used a Wilcoxon non-parametric test for trend. We used the Kaplan-Meier survival estimator to depict the time to initial and final viral culture stratified by N-R use and presence versus absence of viral rebound and compared them using log-rank testing. We defined the date of culture conversion as either: 1) the first swab date in participants with no positive cultures during observation; 2) as the midpoint between the final positive culture and the next negative culture in those who had a culture conversion during observation, or 3) the date of the last study specimen for those with a positive culture on the last study specimen. We assessed the validity of symptom worsening, as defined by an increase in TSS by 3 or more points from a prior date, to detect virologic rebound.[7] Finally, we report the proportion of sequenced viruses before and after the occurrence of virologic rebound with mutations in the NSP5 gene encoding $M^{pro}$ of SARS-CoV-2. Statistical analyses and figure production were conducted with Stata version 16.1 and GraphPad Prism version 9.5.

*Ethical Considerations*

All study participants provided verbal informed consent. Written consent was waived by the review committee based on the need to obtain consent for a minimal risk study during the acute phase of COVID-19 infection. The study procedures were approved by Institutional Review Board and the Institutional Biosafety Committee at Mass General Brigham.

**Supplemental Figure S1.** Screening and enrollment diagram.

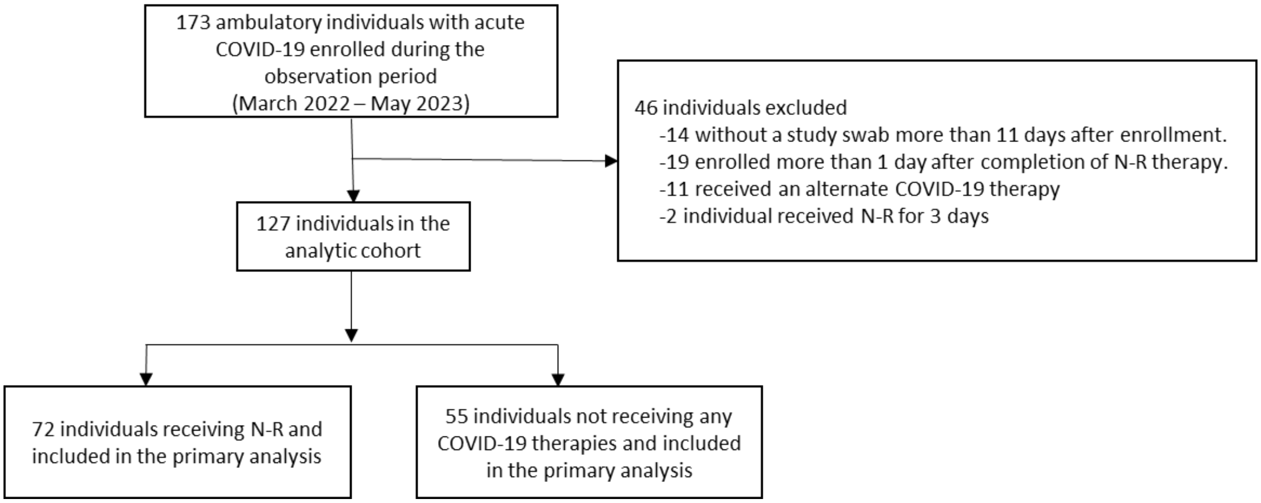

**Supplemental Figure S2.** Sequencing of NSP5 for nirmatrelvir resistance mutations.

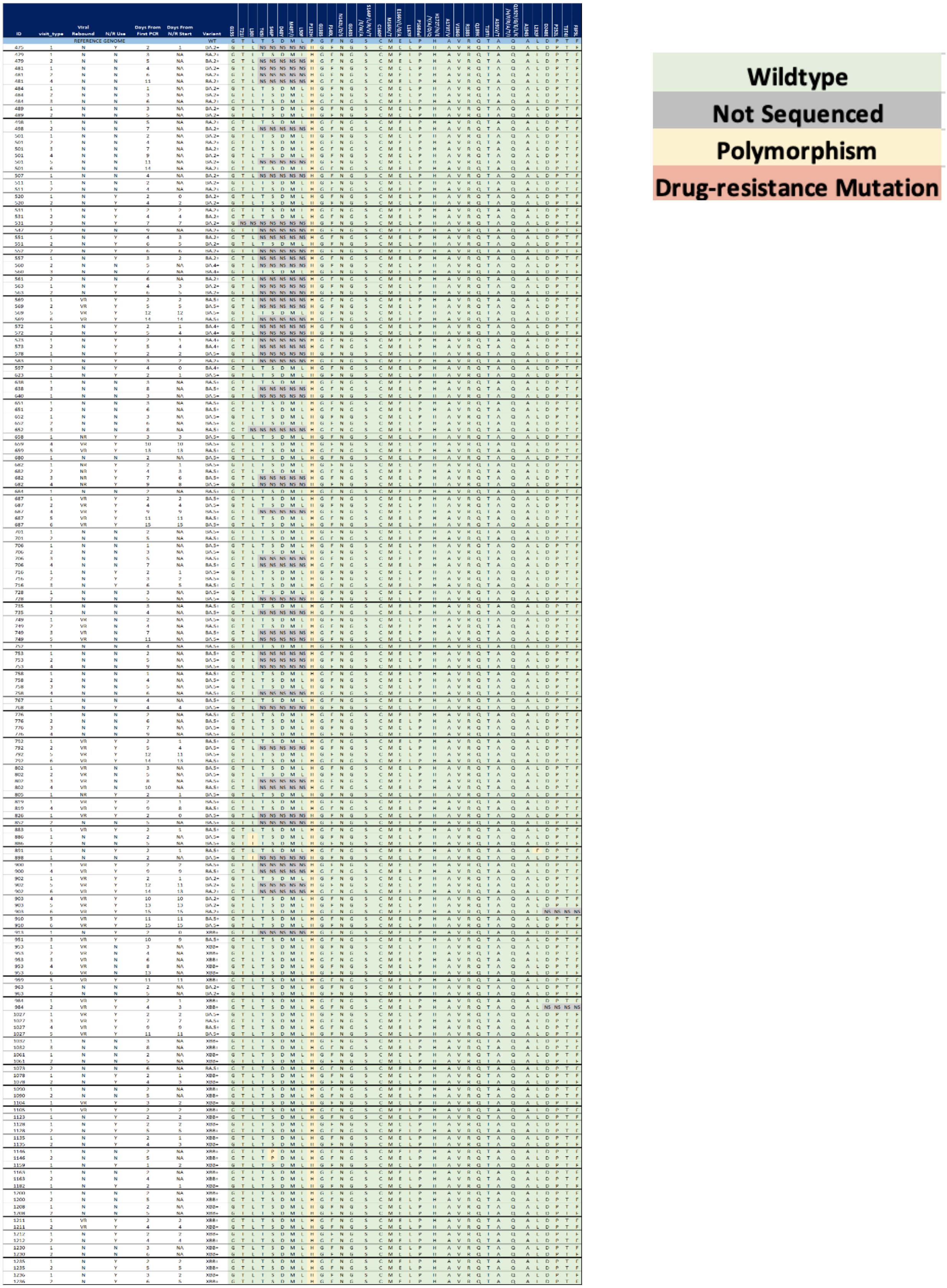

**Supplemental Figure S3.** Individual decay curves for virologic rebounders. The number above each graph corresponds to the participant's study ID. ID 953 (black box) did not receive nirmatrelvir-ritonavir. Abbreviations: TCID50, median tissue culture infectious dose; N/R, nirmatrelvir-ritonavir; IC, immunocompromised (full details are available in Supplemental Table S1).

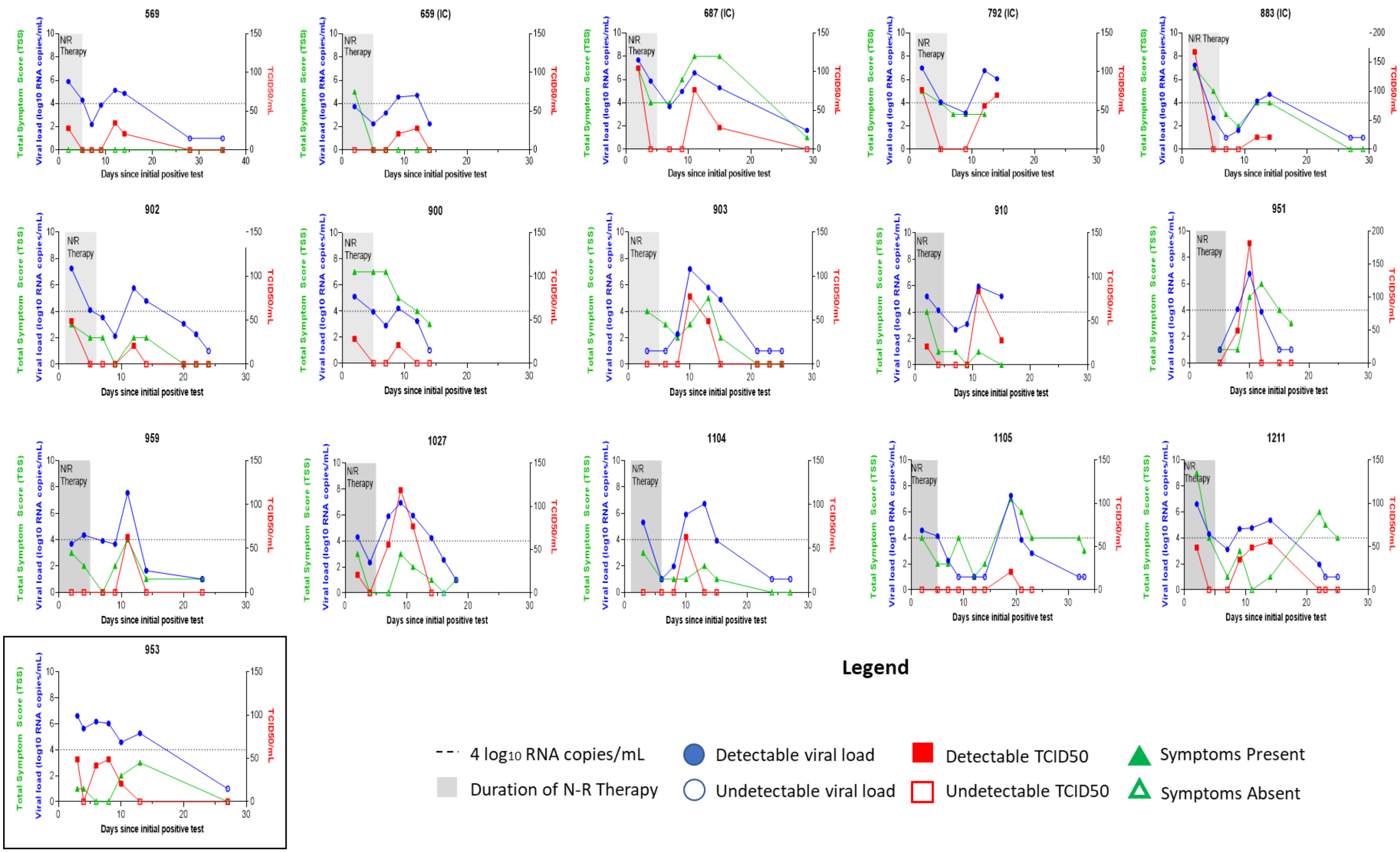

**Supplemental Table S1.** Clinical characteristics of individuals with immunosuppressing conditions or therapies in the cohort.

| ID | Diagnosis | Treatment | COVID-19 Therapy |
|---|---|---|---|
| 475 | Multiple sclerosis | Rituximab within 12 months of COVID-19 | Nirmatrelvir-Ritonavir |
| 531 | Sarcoidosis | Infliximab | Nirmatrelvir-Ritonavir |
| 547 | Rheumatoid arthritis | Tocilizumab, Methotrexate | Untreated |
| 549 | Bechet's disease | Azathioprine | Nirmatrelvir-Ritonavir |
| 550 | Rheumatoid arthritis | Methotrexate, Hydroxychloroquine | Nirmatrelvir-Ritonavir |
| 551 | Psoriatic arthritis | Infliximab | Nirmatrelvir-Ritonavir |
| 552 | Seronegative spondyloarthropathy | Adalimumab, Methotrexate | Nirmatrelvir-Ritonavir |
| 557 | Rheumatoid arthritis, systemic lupus erythematosus | Methotrexate | Nirmatrelvir-Ritonavir |
| 563 | Rheumatoid arthritis | Adalimumab | Nirmatrelvir-Ritonavir |
| 569 | Systemic lupus erythematosus | Hydroxychloroquine, Methylprednisolone daily | Nirmatrelvir-Ritonavir |
| 573 | Inflammatory arthritis | Adalimumab, Hydroxychloroquine | Nirmatrelvir-Ritonavir |
| 597 | Giant cell arteritis, polymyalgia rheumatica | Tocilizumab, Prednisone daily | Nirmatrelvir-Ritonavir |
| 658 | Rheumatoid arthritis | Tocilizumab | Nirmatrelvir-Ritonavir |
| 678 | Rheumatoid arthritis | Tofacitinib | Nirmatrelvir-Ritonavir |
| 687 | Systemic lupus erythematosus, rheumatoid arthritis | Hydroxychloroquine, methotrexate | Nirmatrelvir-Ritonavir |
| 691 | Rheumatoid arthritis | Rituximab | Nirmatrelvir-Ritonavir |
| 716 | Rheumatoid arthritis | Methotrexate | Nirmatrelvir-Ritonavir |
| 723 | Multiple sclerosis, acquired hypogammaglobulinemia | IVIG every 4 weeks; Ocrelizumab within 12 months | Untreated |
| 725 | Rheumatoid arthritis | Infliximab, methotrexate, hydroxychloroquine | Nirmatrelvir-Ritonavir |
| 735 | Psoriatic arthritis | Adalimumab | Untreated |
| 768 | Ankylosing spondylitis | Secukinumab | Nirmatrelvir-Ritonavir |
| 805 | Ulcerative colitis, inflammatory arthritis | Golimumab, methotrexate | Nirmatrelvir-Ritonavir |
| 892 | HIV infection | N/A, on antiretroviral therapy, CD4 cell count>200 | Untreated |
| 945 | IgG4 related disease | Rituximab | Nirmatrelvir-Ritonavir |
| 952 | Inflammatory arthritis | Adalimumab | Nirmatrelvir-Ritonavir |
| 953 | Rheumatoid arthritis | Infliximab, methotrexate, prednisone | Untreated |
| 1235 | Psoriatic arthritis | Etanercept, methotrexate | Nirmatrelvir-Ritonavir |
| 1236 | Systemic lupus erythematosus | Belimumab, methotrexate, prednisone | Nirmatrelvir-Ritonavir |

**Supplemental Table S2.** Cohort characteristics.

| Characteristic | Receipt of Nirmatrelvir-Ritonavir (n=72) | No Receipt of Nirmatrelvir-Ritonavir (n=55) | P-value |
|---|---|---|---|
| Age (median/IQR) | 57 (46-71) | 39 (31-57) | 0.001 |
| Gender (n, %) | | | 1.00 |
| Female | 54 (75) | 42 (76) | |
| Male | 18 (25) | 13 (24) | |
| Race (n, %) | | | 0.83 |
| White | 57 (79) | 40 (73) | |
| Black/AA | 7 (10) | 5 (9) | |
| Asian | 2 (3) | 3 (5) | |
| Other | 3 (4) | 4 (7) | |
| Unknown | 3 (4) | 3 (6) | |
| Ethnicity (n, %) | | | 0.09 |
| Hispanic/Latino | 6 (8) | 4 (7) | |
| Non-Hispanic/Latino | 62 (86) | 41 (75) | |
| Other/Unknown | 4 (6) | 10 (18) | |
| COVID-19 Vaccines (median/IQR) | 4 (3-5) | 3 (3-4) | 0.001 |
| Days since last vaccine (median/IQR) | 132 (75-253) | 185 (133-315) | 0.017 |
| Immunosuppression[a] (n, %) | | | 0.002 |
| Absent | 49 (68) | 50 (91) | |
| Present | 23 (32) | 5 (9) | |
| COVID-19 Variant (n, %) | | | 0.63 |
| BA.2[b] | 10 (14) | 10 (18) | |
| BA.5[c] | 19 (26) | 20 (36) | |
| XBB[d] | 15 (21) | 9 (16) | |
| Other | 3 (4) | 2 (4) | |
| Incomplete[e] | 25 (35) | 14 (26) | |
| Reason for Baseline Test[f] | | | 0.41 |
| Symptoms | 65 (90) | 45 (82) | |
| Exposure | 6 (8) | 7 (13) | |
| Screening | 1 (2) | 2 (4) | |
| Other | 0 (0) | 1 (1) | |
| Baseline Test Type (n, %) | | | 0.026 |
| PCR | 39 (46) | 41 (75) | |

| | | | |
|---|---|---|---|
| Rapid Antigen | 33 (54) | 14 (25) | |
| Baseline Test Ct Value Available (n, %) | 24 (33) | 32 (58) | |
| Baseline Test Ct Value (median/IQR) | 21.9 (17.2-26.4) | 23.2 (19.7-31.4) | 0.28 |
| Days from Symptom Onset to Baseline Test (median/IQR) | 1 (1-2) | 2 (1-3) | 0.041 |

[a] Immunosuppression defined as presence of an immunosuppressing condition or use of an immunosuppressing medication, as determined by physician chart review. Full details of these conditions are available in Supplemental Table S1.

[b] Includes BA.2 subvariants

[c] Includes BA.5 subvariants

[d] Includes XBB subvariants

[e] Only genomes with ≥24000 base pair sequence lengths were considered complete

[f] Participants could select multiple reasons for testing. We categorized them such that symptoms take precedence, followed by exposure, and then followed by screening.

**Supplemental Table S3**: Logistic regression model of correlates of virologic rebound with acute COVID-19

| | Univariable Models | | Multivariable Models | |
|---|---|---|---|---|
| **Characteristic** | **OR (95%CI)** | **P-value** | **AOR (95%CI)** | **P-value** |
| Age | | | | |
| <50 | REF | | REF | |
| ≥50 | 4.10 (1.11-15.21) | 0.035 | 1.50 (0.34-6.62) | 0.59 |
| Sex | | | | |
| Male | REF | | REF | |
| Female | 0.48 (0.10-0.59) | 0.19 | 0.51 (0.15-1.72) | 0.28 |
| Vaccinations | | | | |
| <3 | REF | | REF | |
| ≥3 | 6.40 (1.39-29.47) | 0.017 | 3.05 (0.61-15.37) | 0.18 |
| Immunosuppression | | | | |
| Absent | REF | | REF | |
| Present | 0.79 (0.21-3.01) | 0.73 | 0.55 (0.13-2.33) | 0.42 |
| N-R Use | | | | |
| Not treated | REF | | REF | |
| N-R treated | 14.21 (1.81-111.29) | 0.011 | 10.02 (1.13-88.74) | 0.038 |

OR: Odds ratio; AOR: adjusted odds ratio; N-R: nirmatrelvir-ritonavir

**Supplemental Table S4.** Median number of days to first and final culture conversion.

| | Median (IQR) days to first negative viral culture | P-value (compared to no therapy group) | Median (IQR) days to final negative viral culture | P-value (compared to no therapy group) |
|---|---|---|---|---|
| No therapy group | 4 (3-6) | REF | 4 (3-6) | REF |
| All N/R users | 3 (2-4) | <0.001 | 4 (2-6) | 0.294 |
| N/R rebound | 3 (3-4) | 0.022 | 14 (13-20)* | <0.001 |
| N/R no rebound | 3 (2-4) | <0.001 | 3 (2-4) | <0.001 |

* Two participants with virologic rebound were culture-positive at their last study timepoint

**Supplemental Table S5.** Virologic characteristics of individuals experiencing virologic rebound.

Rebound after nirmatrelvir-ritonavir use

| ID | Initial viral load nadir ($log_{10}$ RNA copies/mL)[a] | Days to initial nadir* | Days to detection of virologic rebound* | Days from end of N-R therapy to detection of rebound* | Viral load peak during rebound ($log_{10}$ RNA copies/mL) | Culturable virus during rebound | Any symptoms during rebound | Symptom rebound (TSS ≥3) | Days to final negative viral load* | Days to final negative viral culture* |
|---|---|---|---|---|---|---|---|---|---|---|
| 569 | 2.2 | 7 | 12 | 7 | 5.1 | Yes | No | No | 21 | 21 |
| 659 | 2.3 | 6 | 10 | 6 | 4.7 | Yes | No | No | 15[b] | 14 |
| 687 | 3.7 | 7 | 9 | 4 | 6.6 | Yes | Yes | Yes | 29[b] | 22 |
| 792 | 3.1 | 9 | 12 | 6 | 6.8 | Yes | Yes | No | 14[b] | 14[b] |
| 883 | 1.0 | 7 | 12 | 6 | 4.7 | Yes | Yes | No | 21 | 21 |
| 900 | 2.9 | 7 | 9 | 4 | 4.2 | Yes | Yes | No | 13 | 11 |
| 902 | 2.1 | 9 | 12 | 6 | 5.8 | Yes | Yes | No | 23 | 13 |
| 903 | 1.0 | 3 | 10 | 5 | 7.2 | Yes | Yes | Yes | 18 | 14 |
| 910 | 2.7 | 7 | 11 | 6 | 6.0 | Yes | Yes | No | 15[b] | 15[b] |
| 951 | 1.0 | 5 | 8 | 2 | 6.8 | Yes | Yes | Yes | 14 | 11 |
| 959 | 3.7 | 9 | 11 | 6 | 7.6 | Yes | Yes | Yes | 19[b] | 13 |
| 1027 | 2.3 | 4 | 7 | 2 | 6.9 | Yes | Yes | Yes | 17 | 13 |
| 1104 | 1.0 | 6 | 10 | 4 | 6.7 | Yes | Yes | No | 20 | 12 |
| 1105 | 1.0 | 9 | 19 | 14 | 7.3 | Yes | Yes | Yes | 28 | 20 |
| 1211 | 3.1 | 7 | 9 | 4 | 5.4 | Yes | Yes | Yes | 23 | 18 |
| | Median/IQR | Median/IQR | Median/IQR | Median/IQR | Median/IQR | n, % | n, % | n, % | Median/IQR | Median/IQR |
| | 2.3 (1.0-3.1) | 7 (6-9) | 10 (9-12) | 6 (4-6) | 6.6 (5.1-6.9) | 15 (100%) | 13 (87%) | 7 (47%) | 19 (15-23) | 14 (13-20) |

Rebound after no therapy

| ID | Initial viral load nadir ($log_{10}$ RNA copies/mL)[a] | Days to initial nadir* | Days to detection of virologic rebound* | | Viral load peak during rebound ($log_{10}$ RNA copies/mL) | Culturable virus during rebound | Any symptoms during rebound | Symptom rebound (TSS ≥3) | Days to final negative viral load* | Days to final negative viral culture* |
|---|---|---|---|---|---|---|---|---|---|---|
| 953 | 5.6 | 4 | 6 | | 6.2 | Yes | Yes | Yes | 20 | 12 |

* Days are from initial positive PCR

[a] Undetectable viral loads imputed as 1.0 $log_{10}$ RNA copies/mL

[b] Final study specimen with detectable viral load or positive viral culture

**Supplemental Table S6.** Validity of symptom rebound to detect virologic rebound.

| | Symptomatic Rebound | No Symptomatic Rebound | Total |
|---|---|---|---|
| Primary Virologic Rebound | 8 | 8 | 16 |
| No Primary Virologic Rebound | 19 | 92 | 111 |
| Total | 27 | 100 | 127 |

| **Measure** | **Estimate** | **95%CI** |
|---|---|---|
| Sensitivity | 50% (8/16) | 25-75% |
| Positive predictive value | 30% (8/27) | 14-50% |
| Specificity | 83% (92/111) | 75-89% |
| Negative predictive value | 92% (92/100) | 82-96% |
